# Long-term predictions for COVID-19 pandemic dynamics in Ukraine, Austria and Italy

**DOI:** 10.1101/2020.04.08.20058123

**Authors:** Igor Nesteruk

## Abstract

The SIR (susceptible-infected-removed) model, statistical approach to the parameter identification and the official WHO daily data about the confirmed cumulative number of cases were used to make some estimations for the dynamics of the coronavirus pandemic dynamics in Ukraine, Italy and Austria. The volume of the data sets and the influence of the information about the initial stages of the epidemics were discussed in order to have reliable long-time predictions. The final sizes and durations for the pandemic in these countries are estimated.

## Introduction

Here we consider the development of epidemic outbreak in Italy, Ukraine and Austria caused by coronavirus COVID-19 (2019-nCoV) (see e.g., [1]). Some estimations of the epidemic dynamics in these countries can be found in [2-7]. In particular, the final size of the epidemic in Italy was calculated in [5]. Unfortunately, the real number of confirmed cases in Italy already exceeds the saturation level predicted in [5] on March 27, 2020. In this paper we will try to clarify the predictions for Italy, to estimate the final size and the duration of the epidemic in Austria and Ukraine and to discuss the amounts of data needed for accurate long-term predictions. In this paper we will use the SIR model [8–12] and the statistics-based method of parameter identification [11, 12].

### Data

The official data about the accumulated numbers of confirmed COVID-19 cases in Italy *V*_*j*_, Ukraine *V*_*Uj*_ and Austria *V*_*Aj*_ from WHO daily situation reports (numbers 41-78), [1] are presented in Tables 1 and 2. The corresponding moments of time *t*_*j*_ (measured in days) are also shown in these tables. The data sets presented in Table 1 were used only for comparison with corresponding SIR curves. Table 2 was used for calculations, comparisons and verifications of predictions.

**Table 1.** Official cumulative numbers of confirmed cases in Italy, Austria and Ukraine during the initial stage of epidemics used only for comparison with SIR curves, [1]

| Day in February<br>and March, 2020 | Time moments<br>In days $t_j$ | Number of cases<br>in Italy, $V_j$ | Number of cases<br>in Austria, $V_{Aj}$ | Number of cases<br>in Ukraine, $V_{Uj}$ |
| --- | --- | --- | --- | --- |
| 21 | -1 | 9 | 0 | 0 |
| 22 | 0 | 76 | 0 | 0 |
| 23 | 1 | 124 | 0 | 0 |
| 24 | 2 | 229 | 0 | 0 |
| 25 | 3 | 322 | 2 | 0 |
| 26 | 4 | 400 | 2 | 0 |
| 27 | 5 | 650 | 4 | 0 |
| 28 | 6 | 888 | 5 | 0 |
| 29 | 7 | 1128 | 10 | 0 |
| 1 | 8 | 1689 | 10 | 0 |
| 2 | 9 | 2036 | 18 | 0 |
| 3 | 10 | 2502 | 24 | 1 |
| 4 | 11 | 3089 | 37 | 1 |

**Table 2.** Official cumulative numbers of confirmed cases in Italy, Austria and Ukraine used for calculations, comparisons and verifications of predictions, [1]

| Day in March and<br>April, 2020 | Time moments<br>In days $t_j$ | Number of cases<br>in Italy, $V_j$ | Number of cases<br>in Austria, $V_{Aj}$ | Number of cases<br>in Ukraine, $V_{Uj}$ |
| --- | --- | --- | --- | --- |
| 5 | 12 | 3858 | 47 | 1 |
| 6 | 13 | 4636 | 66 | 1 |
| 7 | 14 | 5883 | 104 | 1 |
| 8 | 15 | 7375 | 112 | 1 |
| 9 | 16 | 9172 | 131 | 1 |
| 10 | 17 | 10149 | 182 | 1 |
| 11 | 18 | 12462 | 302 | 1 |
| 12 | 19 | 15113 | 361 | 3 |
| 13 | 20 | 17660 | 504 | 3 |
| 14 | 21 | 21157 | 800 | 3 |
| 15 | 22 | 24747 | 959 | 3 |
| 16 | 23 | 27980 | 1132 | 7 |
| 17 | 24 | 31506 | 1332 | 14 |
| 18 | 25 | 35713 | 1646 | 16 |
| 19 | 26 | 41035 | 1843 | 16 |
| 20 | 27 | 47021 | 2649 | 26 |
| 21 | 28 | 53578 | 3024 | 47 |
| 22 | 29 | 59138 | 3631 | 47 |
| 23 | 30 | 63927 | 4486 | 84 |
| 24 | 31 | 69176 | 5282 | 113 |
| 25 | 32 | 74386 | 5888 | 156 |
| 26 | 33 | 80539 | 7029 | 218 |
| 27 | 34 | 86498 | 7697 | 311 |
| 28 | 35 | 92472 | 8291 | 418 |
| 29 | 36 | 97689 | 8813 | 480 |
| 30 | 37 | 101739 | 9618 | 549 |
| 31 | 38 | 105792 | 10182 | 669 |
| 1 | 39 | 110574 | 10711 | 804 |
| 2 | 40 | 115242 | 11129 | 987 |
| 3 | 41 | 119827 | 11525 | 1096 |
| 4 | 42 | 124632 | 11766 | 1251 |
| 5 | 43 | 128948 | 11983 | 1319 |
| 6 | 44 | 132547 | 12297 | 1462 |

### SIR model

The SIR model for an infectious disease [7-11] relates the number of susceptible persons *S* (persons who are sensitive to the pathogen and **not protected**); the number of infected is *I* (persons who are sick and **spread the infection**; please don’t confuse with the number of still ill persons, so known active cases) and the number of removed *R* (persons who **no longer spread the infection**; this number is the sum of isolated, recovered, dead, and infected people who left the region); *α* and *ρ* are constants.

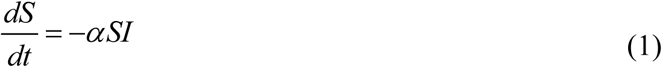

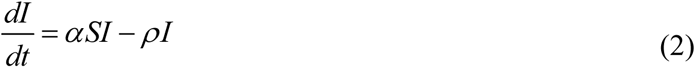

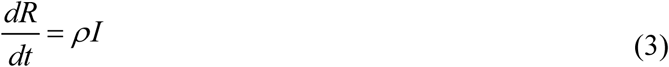

To determine the initial conditions for the set of equations (1–3), let us suppose that at the moment of the epidemic outbreak *t*_0_, [10, 11]:

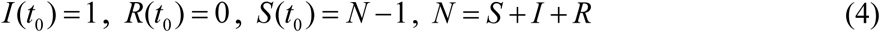

The analytical solution for the set of equations (1–3) was obtained by introducing the function *V* (*t*) = *I* (*t*) + *R*(*t*), corresponding to the number of victims or cumulative confirmed number of cases, [11]:

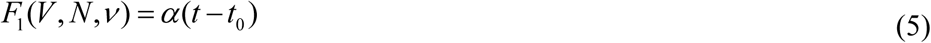

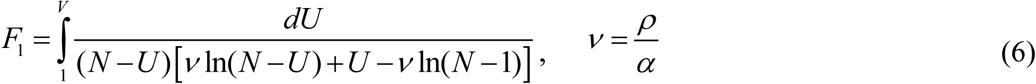

Thus, for every set of parameters *N*, ν, *α, t*_0_ and a fixed value of *V* the integral (6) can be calculated and the corresponding moment of time can be determined from (5). Then functions *I(t)* and *R(t)* can be easily calculated with the of formulas, [11, 12].

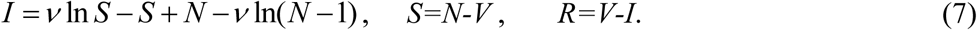

Function *I* has a maximum at *S* = ν and tends to zero at infinity, see [8, 9]. In comparison, the number of susceptible persons at infinity *S*_∞_ > 0, and can be calculated from the non-linear equation, [11, 12]:

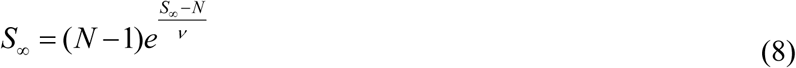

The final number of victims (final accumulated number of cases) can be calculated from:

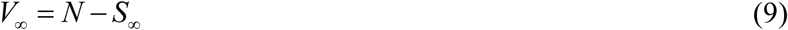

To estimate the duration of an epidemic outbreak, we can use the condition

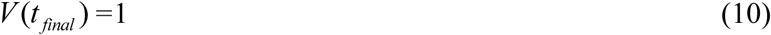

which means that at *t* > *t* _*final*_ less than one person still spread the infection.

### Parameter identification procedure

In the case of a new epidemic, the values of this independent four parameters are unknown and must be identified with the use of limited data sets. A statistical approach was developed in [11] and used in [5, 11, 12, 15] to estimate the values of unknown parameters. The registered points for the number of victims *V*_*j*_ corresponding to the moments of time *t*_*j*_ can be used in order to calculate *F*_1 *j*_ = *F*_1_ (*V*_*j*_, *N*, ν) for every fixed values *N* and ν with the use of (6) and then to check how the registered points fit the straight line (5). For this purpose the linear regression can be used, e.g., [13], and the optimal straight line, minimizing the sum of squared distances between registered and theoretical points, can be defined. Thus we can find the optimal values of *α t*_0_ and calculate the correlation coefficient *r*.

Then the F-test may be applied to check how the null hypothesis that says that the proposed linear relationship (5) fits the data set. The experimental value of the Fisher function can be calculated with the use of the formula:

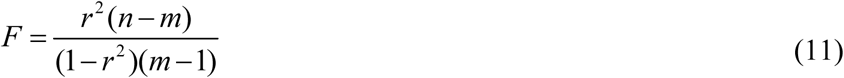

where *n* is the number of observations, *m*=2 is the number of parameters in the regression equation, [13]. The corresponding experimental value *F* has to be compared with the critical value *F*_*C*_ (*k*_1_, *k*_2_) of the Fisher function at a desired significance or confidence level (*k*_1_ = *m* −1, *k*_2_ = *n* − *m*), [14]. When the values *n* and *m* are fixed, the maximum of the Fisher function coincides with the maximum of the correlation coefficient. Therefore, to find the optimal values of parameters *N* and ν, we have to find the maximum of the correlation coefficient. To compare the reliability of different predictions (with different values of *n*) it is useful to use the ratio *F* / *F*_*C*_ (1, *n* − 2) at fixed significance level, [15]. We will use the level 0.001; corresponding values *F*_*C*_ (1, *n* − 2) can be taken from [14]. The most reliable prediction yields the highest *F* / *F*_*C*_ (1, *n* − 2) ratio.

### Results for Italy

The first preliminary prediction for Italy was published in [5] on March 27, 2020. Its results are presented in the first column of Table 3. Usually the number of cases during the initial period of an epidemic outbreak is not reliable. To avoid their influence on the results, only *V*_*j*_ values for the period March 5-22, 2020 (12 ≤ *t* _*j*_ ≤ 29, see Table 2) were used to calculate this first prediction. For other predictions different values of the initial *t*_*j1*_ and final *t*_*j2*_ moments of time were used. They are shown in Tables 3-5 together with corresponding number of observations *n*. The SIR curves for the prediction No. 4 (calculated with the use of the recent *V*_*j*_ values without taking into account maximum number of initial moments of time) are shown in Fig. 1. Markers represent the *V*_*j*_ values taken for calculations (“circles”); for a comparison (“triangles”) and a verification of predictions (“star”).

**Table 3.** Predictions for the epidemic in Italy. Optimal values of parameters and other SIR model characteristics.

| Number of prediction | 1, [5], March 27, 2020 | 2 | 3 | 4 |
| --- | --- | --- | --- | --- |
| $N$ | 534656 | 669312 | 723763.2 | 738240 |
| $\nu$ | 476712.7441113 | 598876.49226424 | 646709.93100256 | 658709.4638592 |
| $\alpha$ | 3.47467962e-06 | 2.589545071e-06 | 2.2076760085e-06 | 2.0049693740e-06 |
| $t_0$ | -18.3797687613 | -22.039683744 | -25.171859952631 | -28.810667619253 |
| $\rho$ | 1.656424058815 | 1.5508176688225 | 1.4277259991363 | 1.3206923014088 |
| $1/\rho$ | 0.603710139730 | 0.6448211289462 | 0.7004145057279 | 0.7571786395160 |
| $t_{j1}$ | 12 | 17 | 17 | 23 |
| $t_{j2}$ | 29 | 37 | 43 | 43 |
| $n$ | 18 | 21 | 27 | 21 |
| $r$ | 0.999378228510 | 0.999725363341 | 0.9992432785563 | 0.9992024895 |
| $F$, eq. (11) | 12854.46446586 | 34576.908427910 | 16499.880328838 | 11897.820763752 |
| $F_C(1, n-2)$ | 16.2 | 15.2 | 13.9 | 15.2 |
| $F/F_C(1, n-2)$ | 793.4854608556 | 2274.7966070993 | 1187.0417502761 | 782.75136603635 |
| $S_\infty$, eq (8) | 423108.1863318 | 533557.58021716 | 575322.57589364 | 585099.75620840 |
| $V_\infty$, eq (9) | 111547.8136681 | 135754.419782840 | 148440.624106361 | 153140.243791598 |
| $t_{final}$, eq (10) | 79.5 | 87.8 | 93.9 | 98.8 |

**Fig. 1.**
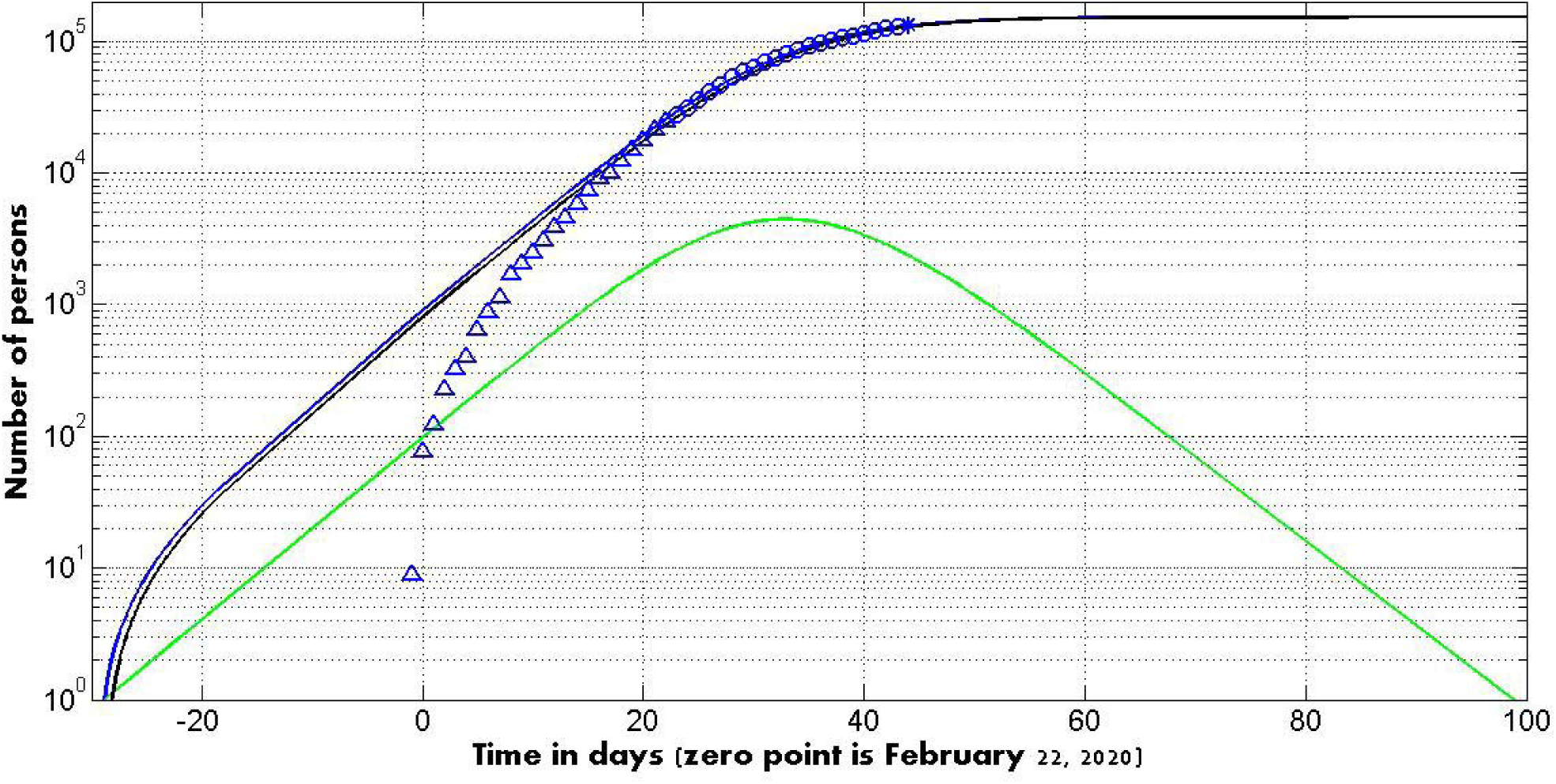
Italy: SIR curves (lines) and accumulated number of cases (markers) versus time. Numbers of infected *I* (green), removed *R* (black) and the number of victims *V*=*I*+*R* (blue line).

It can be seen that the real epidemic outbreak could start even in January 2020. Probably, first cases of COVID-2019 infection were not identified and sick people were not isolated. It could be a reason of the high saturation level of the epidemic (approximately 148,000 −153,000 according to the last predictions 3 and 4). The new cases could stop to appear at the moments of time 93 −99 (see two last values in the last row of Table 3). These moments correspond to May 25-31, 2020. The average time of spreading infection 1/ *ρ* could be estimated as 0.7-0.75 days (according to last two predictions, see Table 3). By comparison, in South Korea was approximately 4.3 hours, [16].

### Results for Austria

For this country the results of calculations are shown in Table 4. Fig. 2 represents the results for the prediction No. 2 corresponding to the highest value of *F* / *F*_*C*_ (1, *n* − 2). It can be seen that first cases of COVID-2019 infection were probably timely identified and sick people were isolated. This may be a reason for much lower saturation level of the epidemic in Austria (approximately 12,000 −13,000 according to the last predictions 2 and 3). The new cases could stop to appear at the moments of time 62 −65 (see two last values in the last row of Table 4). These moments correspond to April 24-27, 2020. The average time of spreading infection 1/ *ρ* could be estimated as 0.44-0.47 days and is much lower than in Italy, but higher than in South Korea, [16].

**Table 4.** Predictions for the epidemic in Austria. Optimal values of parameters and other SIR model characteristics.

| Number of prediction | 1 | 2 | 3 |
| --- | --- | --- | --- |
| $N$ | 71099.2204800000 | 65613.6960000000 | 64200 |
| $\nu$ | 63712.2688217289 | 58821.6712255282 | 57523.2000000000 |
| $\alpha$ | 3.190277719e-05 | 3.63707317842e-05 | 3.91631629710e-05 |
| $t_0$ | 2.38559451409778 | 3.38412182748648 | 4.88499029942458 |
| $\rho$ | 2.03259831679520 | 2.13938722724589 | 2.25279045621570 |
| $1/\rho$ | 0.491981121767681 | 0.467423562814918 | 0.443893925971183 |
| $t_{j1}$ | 21 | 21 | 28 |
| $t_{j2}$ | 36 | 43 | 43 |
| $n$ | 16 | 23 | 16 |
| $r$ | 0.998063248602538 | 0.999267711104992 | 0.999476595767646 |
| $F$ | 3603.80309527417 | 14322.8552686895 | 13363.4852428387 |
| $F_C(1, n-2)$ | 17.3000000000000 | 14.6000000000000 | 17.3000000000000 |
| $F/F_C(1, n-2)$ | 208.312317645906 | 981.017484156815 | 772.455794383737 |
| $S_\infty$ | 56848.1710083937 | 52507.9044514993 | 51318.7804944884 |
| $V_\infty$ | 14251.0494716063 | 13105.7915485007 | 12881.2195055116 |
| $t_{final}$ | 67.3 | 64.5 | 62.6 |

**Fig. 2.**
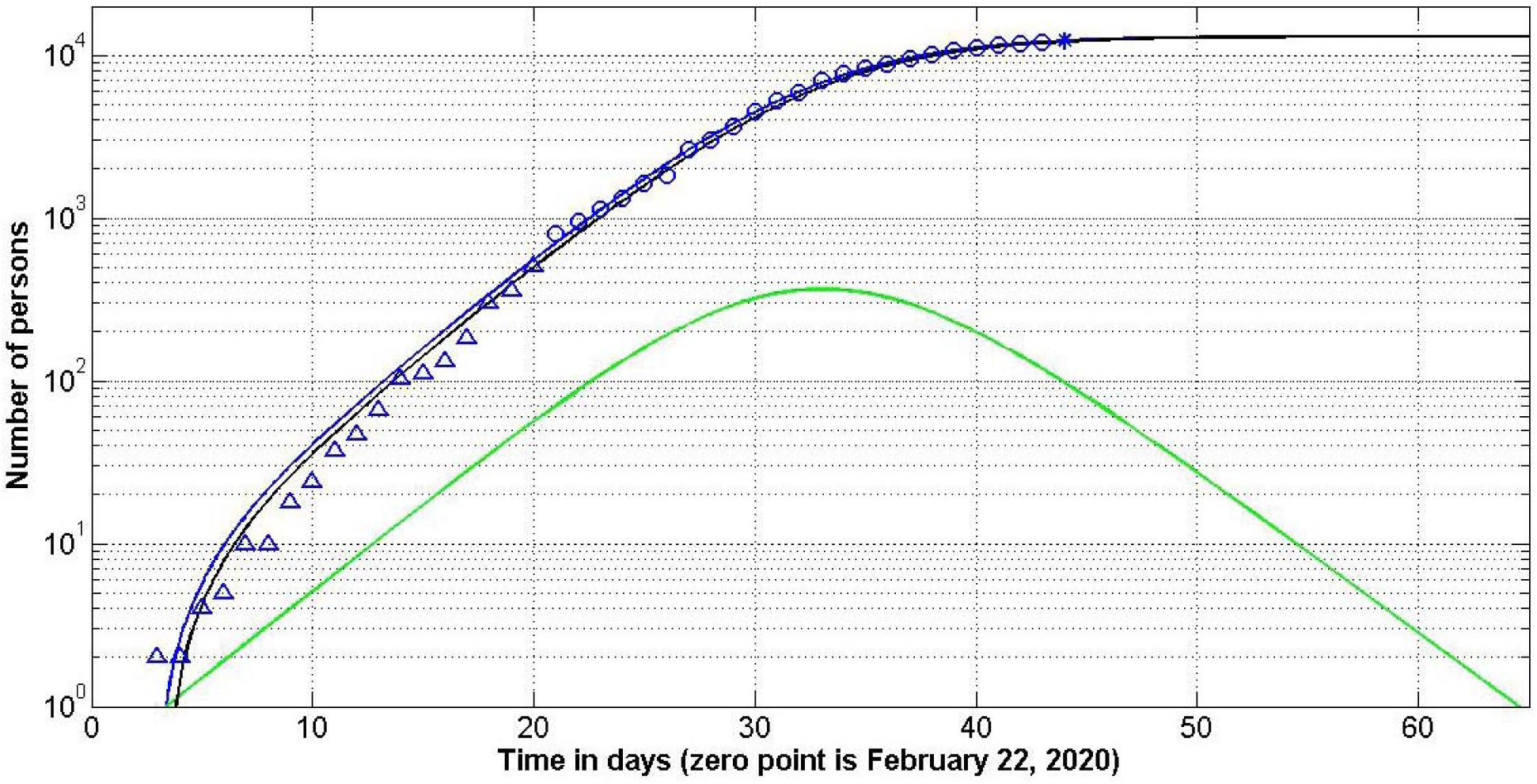
Austria: SIR curves (lines) and accumulated number of cases (markers) versus time. Numbers of infected *I* (green), removed *R* (black) and the number of victims *V*=*I*+*R* (blue line); “circles” show the cases taken for calculations; “triangles” correspond to the cases during initial stage of the epidemic; “star” –the last data point used only for a verification of the prediction.

### Results for Ukraine

The results of calculations are shown in Table 5. Fig. 3 represents the results for the prediction No. 6-1 corresponding to highest number of initial points which were not used for calculations. Since the epidemic outbreak occurred later than in Italy and Austria, the number of observations in Ukraine is lower and presented predictions have to be treated as very preliminary ones. It can be seen that the first case of COVID-2019 infection in Chernivtsi was probably timely identified and isolated. The epidemic outbreak was probably caused by other persons. Preliminary estimations of the saturation level are approximately 1,700 −2,000 (according to the last predictions 2-2, 5-2 and 6-1). The new cases could stop to appear at the moments of time 57-61 (see three last values in the last row of Table 5). These moments correspond to April 19-23, 2020. The average time of spreading infection 1/ *ρ* could be estimated as 0.36-0.4 days.

**Table 5.**
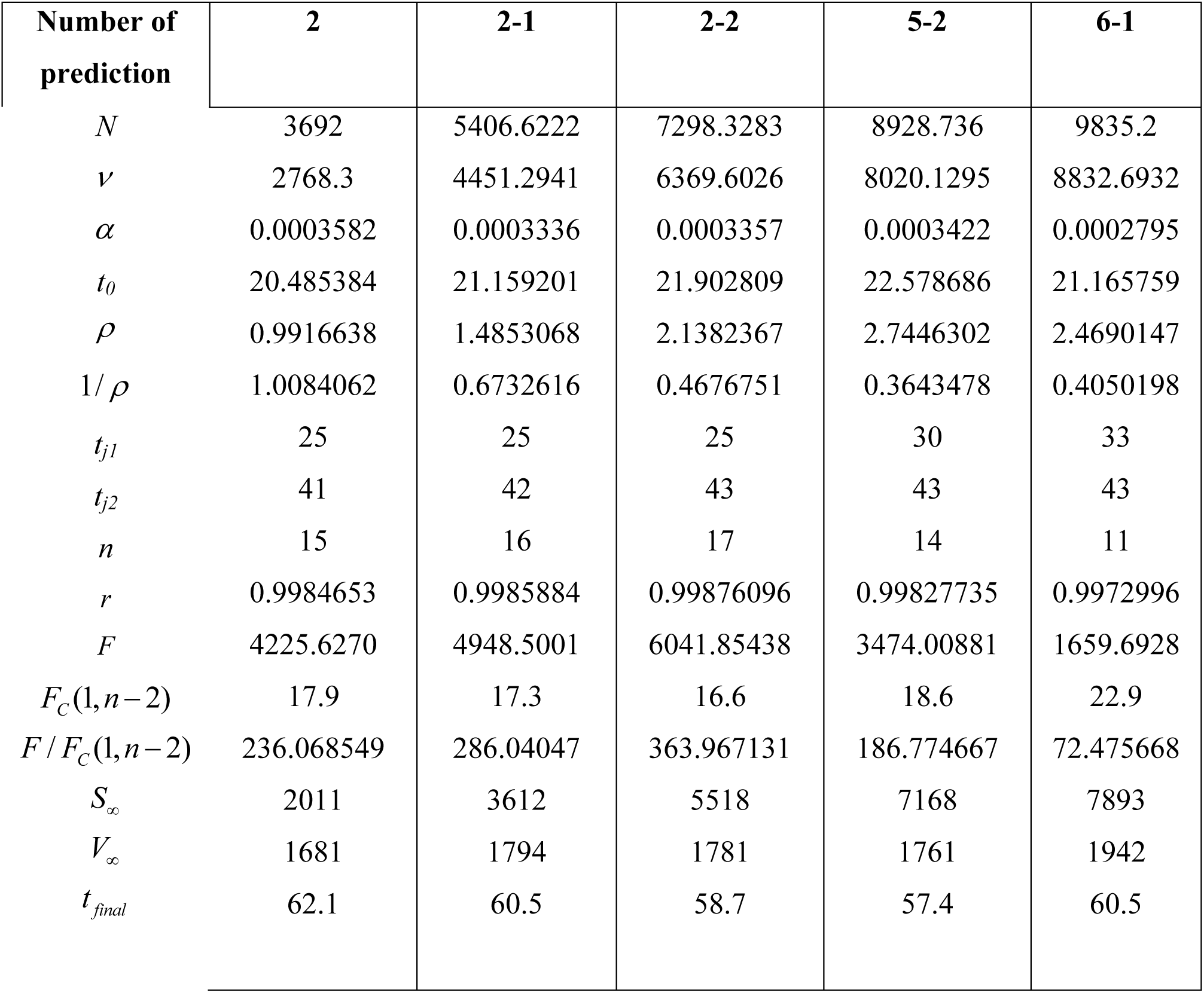
Predictions for the epidemic in Ukraine. Optimal values of parameters and other SIR model characteristics.

**Fig. 3.**
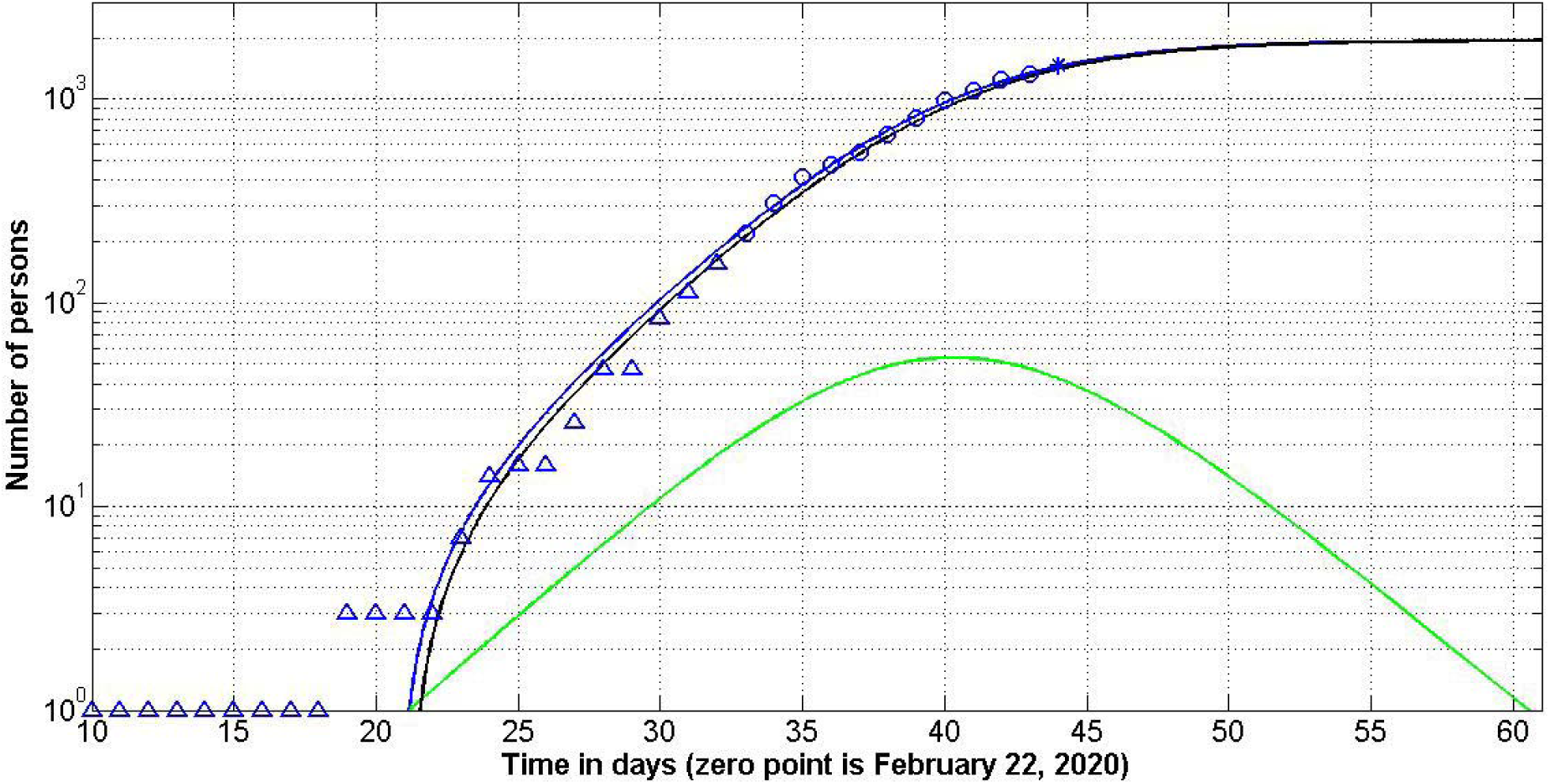
Ukraine: SIR curves (lines) and accumulated number of cases (markers) versus time. Numbers of infected *I* (green), removed *R* (black) and the number of victims *V*=*I*+*R* (blue line); “circles” show the cases taken for calculations; “triangles” correspond to the cases during initial stage of the epidemic; “star” –last data point used only for a verification of the prediction.

## Discussion

The accuracy of any mathematical model is limited. The used SIR model is not an exception. The real processes are much more complicated. In particular, all the parameters in SIR model are supposed to be constant. If the quarantine measures and speed of isolation change or new infected persons are coming in the country, the accuracy of the prediction reduces. The accuracy of predictions increases with increasing the number of observations. On the other hand, the need for forecasts is reduced if an epidemic is stabilized.

## Conclusions

The SIR (susceptible-infected-removed) model and statistical approach to the parameter are able to make some reliable estimations for the epidemic outbreaks. The accuracy of long-term predictions is limited by uncertain information, especially at the beginning of an epidemic. The long enough observations may eliminate the influence of the initial stage data and increase the accuracy of predictions. Even at limited amount of data the SIR model can be used to estimate the final size of an epidemic and its duration. The further course of COVID-19 pandemic in Ukraine, Austria and Italy will show the real accuracy of the proposed method.

## Data Availability

data are in text

## Acknowledgements

I would like to express my sincere thanks to Gerhard Demelmair and Ihor Kudybyn for their help in collecting and processing data.

## Notes

### Competing Interest Statement

The authors have declared no competing interest.

### Funding Statement

no funding

